# Performance evaluation of the point-of-care SAMBA II SARS-CoV-2 Test for detection of SARS-CoV-2

**DOI:** 10.1101/2020.05.24.20100990

**Authors:** Sonny M Assennato, Allyson V Ritchie, Cesar Nadala, Neha Goel, Hongyi Zhang, Rawlings Datir, Ravindra K Gupta, Martin D Curran, Helen H Lee

**Affiliations:** Diagnostics for the Real World EU Ltd., Chesterford Research Park, Great Chesterford, UK; Diagnostics for the Real World Ltd., San Jose, USA; Clinical Microbiology and Public Health Laboratory, PHE Cambridge, Addenbrooke’s Hospital, Cambridge, UK; Division of Infection and Immunity, University College London, London, UK; Department of Medicine, University of Cambridge, Cambridge, UK; Africa Health Research Institute, Durban, South Africa

**Keywords:** SAMBA II, SARS-CoV-2, point-of-care, COVID-19

## Abstract

Nucleic acid amplification for the detection of SARS-CoV-2 RNA in respiratory samples is the standard method for diagnosis. These tests are centralised and therefore turnaround times can be 2-5 days. Point-of-care testing with rapid turnaround times would allow more effective triage in settings where patient management and infection control decisions need to be made rapidly.

Inclusivity and specificity of the SAMBA II SARS-CoV-2 assay was determined by *in silico* analyses of the primers and probes. Analytical and clinical sensitivity and specificity of the SAMBA II SARS-CoV-2 Test was evaluated for analytical sensitivity and specificity. Clinical performance was evaluated in residual clinical samples compared to the Public Health England reference tests.

The limit of detection of the SAMBA II SARS-CoV-2 Test is 250 cp/mL and is specific for detection of 2 regions of the SARS-CoV-2 genome. The clinical sensitivity was evaluated in 172 clinical samples provided by the Clinical Microbiology and Public Health Laboratory, Addenbrooke’s Hospital, Cambridge (CMPHL), which showed a sensitivity of 98.9% (95% CI 94.03-99.97%), specificity of 100% (95% CI 95.55-100%), PPV of 100% and NPV of 98.78% (92.02-99.82%) compared to testing by CMPHLSAMBA detected 3 positive samples that were initially negative by PHE Test. The data shows that the SAMBA II SARS-CoV-2 Test performs equivalently to the centralised testing methods with a much quicker turnaround time. Point of care testing, such as SAMBA, should enable rapid patient management and effective implementation of infection control measures.

## Introduction

The SARS-CoV-2 was first reported in Wuhan, China, in early December 2019 and is the causative agent of coronavirus disease 19 (COVID-19) [1]. It has since spread to over 188 countries/regions around the world [2], causing 317,234 deaths [3]. It was declared a pandemic by the World Health Organisation on the 11th of March 2020 [4]. In Europe, the country with the highest number of deaths is UK, which as of the 17^th^ of May has had 243,694 lab-confirmed cases and 34,636 deaths in all settings [5]. Nucleic acid testing is essential for early diagnosis of SARS-CoV-2 infection as antibody response is often not detected until 7-10 post onset of symptoms [6]. Upper respiratory tract (URT) specimens such as nose and throat swabs generally have high SARS-CoV-2 viral loads upon symptom onset [7]. The standard diagnostic test for SARS-CoV-2 in the UK is done by real-time RT-PCR of the RdRp gene [8], from a combined throat and nose swab sample. The UK has dramatically scaled up testing from 5,000 tests per day in March 2020 to 100,000 tests per day by the end of April. Although this test has good accuracy, the samples must be transported to centralized testing laboratories and batched for processing, which leads to turnaround times of around 48 hours or more. This means treatment of severely ill patients may be suboptimal when other causative pathogens are in the differential diagnosis and those requiring admission or triage with possible COVID-19 maybe unnecessarily isolated or inappropriately cohorted in a COVID-19 ward. This causes obvious bottlenecks in addition to the sheer number of samples that require processing; at present hospital and regional laboratories are at full capacity and a rapid point of care (POC) testing is required.

The SAMBA II nucleic acid testing system was originally designed for HIV testing in POC and resource-limited settings, with CE-marked products for early infant diagnosis [9,10] and viral load monitoring [11-13]. Since 2017, SAMBA HIV tests have been implemented in Uganda, Malawi, Zimbabwe and Central African Republic. The SAMBA II SARS-CoV-2 Test system has now been developed to specifically detect the presence of the novel coronavirus, SARS-CoV-2, in nose and throat swab samples run on the SAMBA II instrument. Test results are available in approximately 1.5 hours. We have here assessed the analytical and clinical performance of the SAMBA II SARS-CoV-2 Test using panels and clinical samples.

## Materials and Methods

### SAMBA sample preparation, amplification, and detection

For the SAMBA II system, sample preparation, amplification, and detection as well as reading and interpretation of the results are fully automated. The test is carried out in the SAMBA II Assay Module under the control of the SAMBA II Tablet Module as previously described for HIV [9-12], according to the manufacturer’s instructions for use. Nose and throat swab samples are preferably resuspended in 2 mL of SAMBA SCoV buffer but the system can be used with viral transport medium (VTM) if the sample is diluted 1:2 with SCoV buffer prior to processing. The input volume for the SAMBA test is 300 μl of which 250 μl is used by the SAMBA II machine as input into the sample preparation. The SAMBA II SARS-CoV-2 Test specifically amplifies and detects two regions of the SARS-CoV-2 genome in the ORF1ab (region 1) and nucleocapsid protein (region 2), reported as two distinct lines on the test strip. A third line on the test strip, the internal control, is present to control for false negatives caused by instrument/reagent problems or inhibition. The presence of one or both test lines indicates a positive result. The presence of just the internal control line indicates a negative result.

### Virus inactivation in SAMBA SCoV buffer

SAMBA SCoV buffer, containing Triton x100, is provided with the SAMBA II SARS-CoV-2 Test for sample collection of the nose and throat swabs. Single round of VSV-G pseudotyped lentiviruses were produced by transfecting HEK-293T cells in a 3-plasmid transfection system (HIV Gag-pol expresser under a CMV promoter, luciferase genome reporter and VSV-g envelope) as previously described [14]. Virus supernatant was harvested after 24 hours and filtered. Fifty (50) μL of pseudovirus preparation was added to increasing dilutions of SAMBA SCoV Buffer in duplicates and incubated at room temperature for 5 minutes. 100μL of the virus-reagent prep was then used to infect 1.0×10^6^TZMbl target cells per well in a 96-well plate and incubated at 37°C, 5% CO2. Forty-eight (48) hours post-infection, luciferase expression was measured using SteadyGlo and a Glomax Luminometer (both Promega).

### *In silico* inclusivity analysis

The SAMBA-SARS-CoV-2 primers and probes for Orf1ab and N regions were individually evaluated using *in-silico* analysis with respect to 157 SARS-CoV-2 sequences in the NCBI database.

### *In silico* specificity analysis

*In silico* analysis for possible cross-reactions with related human Coronaviruses (Human coronavirus 229E, Human coronavirus OC43, Human coronavirus HKU1, Human coronavirus NL63, SARS-coronavirus and MERS-coronavirus) was conducted by mapping primers in the SAMBA II SARS-CoV-2 Test individually to the sequences downloaded from the NCBI database.

An *in silico* analysis for possible cross-reactions with other high-priority organisms was conducted by carrying out a blastn search for each of the SAMBA primers and probes against NCBI databases and retrieving all sequences with homologies > 80%.

### Panels and samples

Panel members for determination of the limit of detection (LOD) were prepared by making serial dilutions of SARS-CoV-2 RNA from strain 2019-nCoV/Italy-INMI1 from EVAg (code number: 008N-03894) in pooled negative combined nose and throat swab samples to target concentrations of 750, 500, 250, 200, 150 and 100 copies/mL.

A Coronavirus RNA specificity panel containing hCoV-NL63, hCoV-229E, hCoV-OC43, MERS-CoV and SARS-CoV HKU339849 were sourced from EVAg (code number 011N-03868). RNA samples from this panel were tested at >100,000 cp/test in the SAMBA II test to determine specificity against other human Coronaviruses.

### Contrived clinical samples

Combined nose and throat swabs were collected from 35 presumed negative individuals using FLOQSwabs (Copan, Italy) and SAMBA SCoV buffer. Thirty contrived positive clinical samples were prepared by spiking known concentrations of SARS-CoV-2 RNA strain 2019-nCoV/Italy-INMI1 (EVAg, Italy) into individual negative specimens to produce final concentrations of 1x LoD (n=3), 2x LoD (n=17), 3x LoD (n=5), 5x LoD (n=3) and 100x LoD (n=2).

### Clinical evaluation

The clinical performance of the SAMBA II SARS-CoV-2 Test was further evaluated retrospectively with 172 residual blinded combined nose and throat swab samples from CMPHL Cambridge laboratory. These residual samples were from symptomatic individuals with suspected Covid-19 from around the East of England region sent for routine laboratory diagnosis and provided as VTM diluted 1:2 with SAMBA SCoV buffer. In total 172 samples were tested by the SAMBA II SARS-CoV-2 Test and results were compared to the Cambridge RdRp gene (Wuhan) assay on the Rotor gene Q real-time PCR assay routinely used by CMPHL based on the publication by Sridhar et al., 2020 [15]. Results are expressed as positive or negative with a Ct cut off of 36 for positive results. Samples were also tested with the PHE Colindale (Reference Laboratory) assay, based on the publication by Corman et al, 2020 [8], which amplifies a different region of the RdRp gene.

For LOD, we used SARS-CoV-2 RNA quantified in cp/mL but in the clinical samples, SAMBA II results were compared to real-time PCR results expressed as Ct number (number of cycles). By systematically testing serial dilutions of samples calibrated in cp/mLby the PHE test a correspondence curve was drawn showing for instance that 25 Ct corresponded to 100,000 cp/mL and 30 Ct to 1,000 cp/mL. The curve obtained is added as supplementary information.

### Research Ethics

Surplus samples obtained from patients known to be symptomatic for COVID-19 and submitted to the CMPHL for routine testing were retrieved before being discarded. These samples were rendered anonymous and provided blinded for the purpose of test validation. Public Health England and NHS Research Ethics Committee have permitted the use of residual samples in this manner, strictly for the purpose of diagnostic assay validation [16]. The evaluation was carried out in accordance with The Use of Human Organs and Tissue act [17].

## Results

### Viral inactivation by SAMBA SCoV collection buffer

Due to the highly infectious nature of SARS-CoV-2, a proprietary buffer which has a low pH and a strong detergent was developed for the SAMBA SARS-CoV-2 Test for sample collection. The ability of the SAMBA SCoV Buffer to inactivate pseudoviruses was assessed by incubating pseudoviruses with increasing dilutions of SAMBA SCoV Buffer in duplicates at room temperature for 5 minutes. Forty-eight (48) hours post-infection, luciferase expression was measured. Figure 1 shows the plot of log_10_ relative light units (RLU) of luciferase expression vs concentration of SAMBA SCoV Buffer containing Triton X100. Results show that luciferase activity was insignificant up to 1:128 dilution of the SAMBA SCoV Buffer, indicating that the pseudovirus was inactivated after 5-minute exposure at room temperature (Figure 1). For the SAMBA II Test, a 10-minute soaking of the swab in the SAMBA SCoV Buffer is recommended.

**Figure 1.**
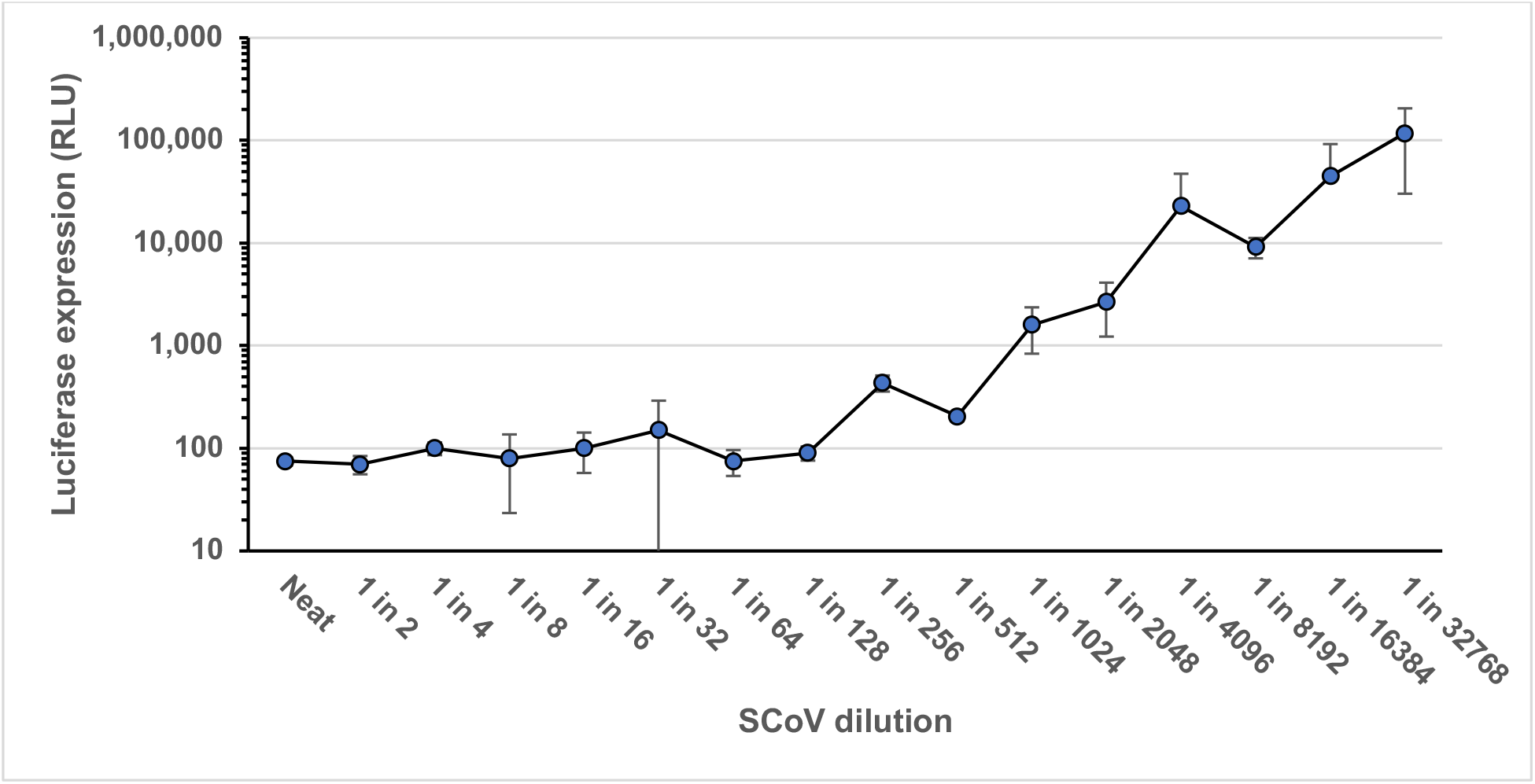
Viral inactivation as demonstrated by Luminescence from a VSV-G pseudotyped reporter virus in descending concentrations of SAMBA Reagent (SCoV Buffer).

### Limit of detection

The limit of detection (LOD) of the SAMBA II SARS-CoV-2 Test was determined using serial dilutions of SARS-CoV-2 RNA in pooled negative combined nose and throat swab samples. The initial LOD was determined by testing 6 levels at target concentrations of 750, 500, 250, 200, 150 and 100 copies/mL. Each panel member was tested in replicates of 3 (Table 1). The final LOD was confirmed by testing 250 copies/mL in replicates of 20, of which all were detected. The results are summarized in Table 1. Therefore, the claimed LOD of the SAMBA II SARS-CoV-2 Test is 250 cp/mL.

**Table 1.** LOD of the SAMBA II SARS-CoV-2 Test

| Concentration SARS-CoV-2<br>RNA (cp/mL) | Number<br>tested | Number<br>positive | % positive |
| --- | --- | --- | --- |
| 750 | 3 | 3 | 100 |
| 500 | 3 | 3 | 100 |
| **250** | **20** | **20** | **100** |
| 200 | 3 | 2 | 66.7 |
| 150 | 3 | 2 | 66.7 |
| 100 | 3 | 0 | 0 |

### Inclusivity

The SAMBA II SARS-CoV-2 Test primers and probes for Target 1 (Orf 1ab) had 100% match to all but one available SARS-CoV-2 sequence for this region in the NCBI database (n=157). For this one sequence, a single nucleotide mismatch was found that maps to the capture probe with no predicted impact on the assay performance. The primers and probes for Target 2 (N) had 100% identity to all available SARS-CoV-2 sequences for this region in the NCBI database (n=157).

### Specificity analysis

*In silico* analysis for possible cross-reactions with related human coronaviruses (Human coronavirus 229E, Human coronavirus OC43, Human coronavirus HKU1, Human coronavirus NL63, SARS-coronavirus and MERS-coronavirus) concluded that none of the SAMBA primers had >80% homology to the organisms listed. In addition to *in silico* analysis, the SAMBA II SARS-CoV-2 Test was evaluated for specificity by using >100,000 cp/test of hCoV-NI63, hCoV-229E, hCoV-OC43, MERS-CoV and SARS-CoV RNA as target in the assay. All samples gave negative results in SAMBA showing that both the Orf1ab and N primers and probes in the SAMBA II SARS-CoV-2 were specific.

An *in silico* analysis for possible cross-reactions with other high-priority organisms showed that only one SAMBA probe (N region) had greater than 80% homology (81 %) to one of the high priority organisms (*Pneumocystis jirovecii* [PJP]). This marginal homology would not impact the performance of the test because the other primers and probes have no homology to *P. jirovecii*. This homology would not be able to compete with 100% homology of the SARS-CoV-2 amplicon to the capture probe.

### Contrived clinical specimens

Positive (n=30) and negative (n=35) swab samples were contrived from individual nose and throat swab samples collected from 35 individuals and tested with the SAMBA II SARS-CoV-2 Test. Spiked positive samples were tested at 1× LOD (n=3), 2× LOD (n=17), 3× LOD (n=5), 5× LOD (n=3) and 100× LOD (n=2) by spiking SARS-CoV-2 RNA into individual negative swab samples. All 35 negative samples were negative and all 30 spiked positive samples were positive when tested with the SAMBA II SARS-CoV-2 Test (Table 2). The overall sensitivity was 100% (95%CI: 88.43-100%) and specificity was 100% (95%CI: 90-100%).

**Table 2.** Clinical performance in 65 contrived clinical samples

| **Concentration SARS-CoV-2 RNA**<br>**(cp/mL)** | **Number**<br>**tested** | **Number**<br>**positive** | **% agreement**<br>**(95% CI)** |
| --- | --- | --- | --- |
| 1-2x LOD | 20 | 20 | 100%<br>(86.1-100%) |
| ≥3x LOD | 10 | 10 | 100 %<br>(74.1-100%) |
| Negative | 35 | 0 | 100%<br>(91.5-100%) |

### Clinical evaluation

The clinical performance of the SAMBA II SARS-CoV-2 Test was further evaluated with 172 combined nose and throat samples from symptomatic individuals provided blinded by CMPHL. The samples were provided in Viral Transport Medium (VTM) diluted 1:2 with SAMBA SCoV buffer. After initial testing there were 87 concordant positives, 81 concordant negatives and 4 discrepant results (3 SAMBA positive and one SAMBA negative) compared with the PHE Colindale reference laboratory test (Table 3). The three SAMBA positive samples were repeat positive by SAMBA and on retest by CMPHL they were found to be borderline positive with high CT values for at least one of the target genes on the Colindale or Cambridge (Wuhan) test (Table 4). The one SAMBA negative sample was negative on repeat by SAMBA but was positive by PHE for RdRp using both the Cambridge (Wuhan) and Colindale assays, with Ct values of 28.87 and 31.18 respectively (Table 4). Therefore, there was just 1 discrepant sample after retest, a false negative for SAMBA (Table 5). From this data set the SAMBA II SARS-CoV-2 Test has a sensitivity of 98.9% (95% CI 94.03-99.97%), specificity of 100% (95% CI 95.55-100%), PPV of 100% and NPV of 98.78% (92.02-99.82%) when compared to the PHE reference tests. The one SAMBA false negative gave a high Ct value on the PHE test (>31), suggesting low viral load, and it was diluted (1:2) for SAMBA testing, which may explain the false-negative result.

**Table 3.**
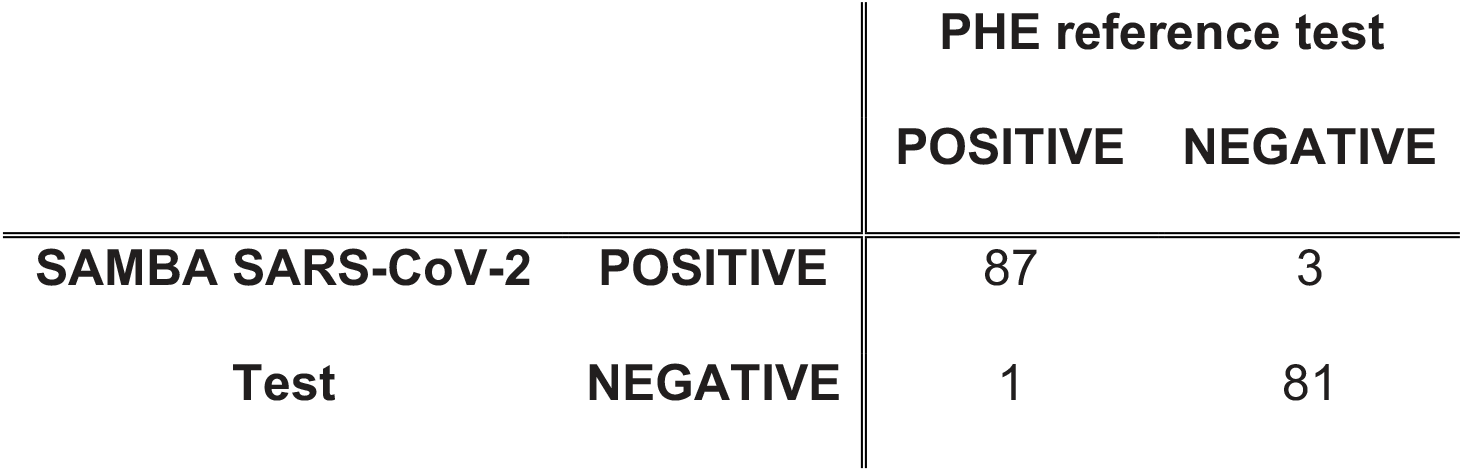
Clinical performance in 172 clinical samples compared to PHE reference test (initial results)

**Table 4.**
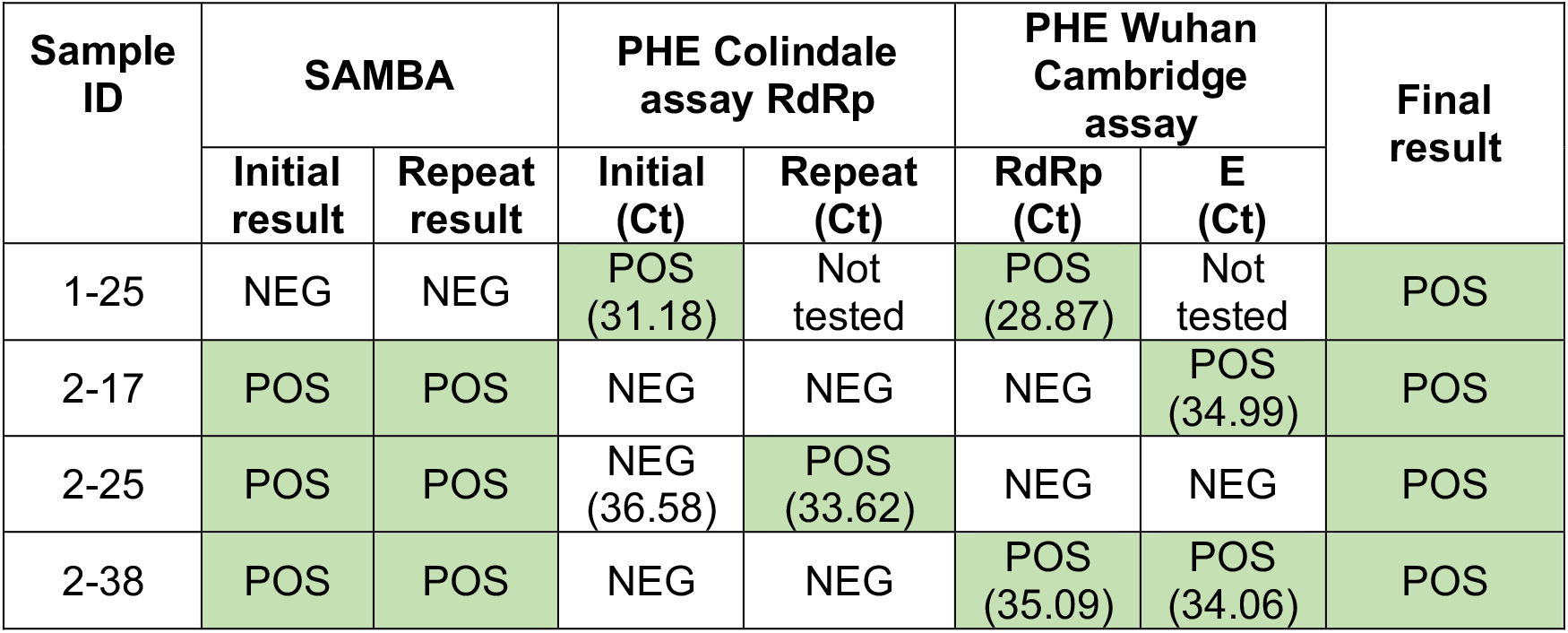
Discrepant analysis of 4 samples

**Table 5.** Clinical performance in 172 clinical samples compared to PHE reference test (initial results)

|  |  |  | PHE reference test |  |
| --- | --- | --- | --- | --- |
|  |  |  | POSITIVE | NEGATIVE |
| SAMBA SARS-CoV-2 Test | POSITIVE |  | 90 | 0 |
|  | NEGATIVE |  | 1 | 81 |

## Discussion

POC molecular tests for SARS-CoV-2, such as SAMBA II, are required to quickly triage patients as centralised testing can take 2-5 days for results. In addition, POC tests would be extremely useful for non-laboratory residential settings such as prisons, immigration centres, nursing homes and rehabilitative centres. The people living in such facilities tend to be vulnerable populations who are at a higher risk for adverse outcome and for infection due to living in close proximity to others [18] and early identification and implementation of increased infection control measures would reduce spread amongst residents and staff. POC would also be extremely useful in other situations where rapid and accurate results are required e.g organ donation, hospital admissions and emergency surgery. POC molecular tests with rapid turnaround, such as SAMBA II, will be essential not only during high rates of infection but also as the country begins to end the lockdown period and localised outbreaks will need to be managed quickly and efficiently.

Our data shows that the SAMBA II SARS-CoV-2 Test is equivalent to centralised testing with excellent sensitivity and specificity. Samples are inactivated in the SAMBA collection buffer and results are available within 86-101 minutes at the POC. High sensitivity and specificity are essential for the appropriate triaging and treatment of incoming patients. The assay has a limit of detection of 250 cp/mL, which is in-line with that claimed by other commercial SARS-CoV-2 tests [19,20]. The specificity of the SAMBA SARS-CoV-2 Test in clinical samples was 100% and the sensitivity was 98.9% compared to the centralised molecular testing by CMPHL. This data includes 3 positive samples detected by SAMBA that were original negative by centralised testing indicating good sensitivity. Clinical evaluation by Zhen et al [21] comparing the performance of Xpert® Xpress SARS-CoV-2 (Cepheid), ePlex® SARS-CoV-2 (GenMark) and ID NOW™ COVID-19 (Abbott) showed limit of detection of 100 cp/mL, 1,000 cp/mL and 10,000 cp/mL and clinical agreement with the reference standard of 98.3%, 91.4% and 87.7% respectively. In another study the Xpert system has been shown to have good agreement with the reference test over a wide range of Ct values, including low positives [22,23] with a positive agreement of 98.9% and negative agreement of 92.0% compared to Roche cobas [22]. The Abbott ID NOW has been shown to have a positive agreement of 73.9% (95% CI: 63.2-82.3%) and negative agreement of 100% (95%CI: 92.9-100%) compared to the lab-based Roche cobas system, with the majority of false negative samples being low viral load (>30 Ct cycles by Roche cobas) [22]. Abbott have since modified the instructions for use to remove the use of swab in transport medium as samples may become too dilute and affect the sensitivity [24].

Potential limitations of this study include that the virus inactivation study was carried out using a constructed pseudovirus rather than a SARS-CoV-2 or other coronavirus due to availability. Also clinical samples were collected in VTM and diluted 1:2 in SCoV buffer rather than collected directly into SCoV buffer, which may affect the sensitivity.

## Data Availability

NA

## Acknowledgements

RKG is funded by Wellcome Senior Fellowship In Clinical Science award no WT108082AIA

